# Prevalence of Abnormal Uterine Bleeding and its Sub-types among Adolescent Girls in India: A Systematic Review and Meta-Analysis

**DOI:** 10.1101/2025.10.24.25338610

**Authors:** Ekta Krishna, Shamshad Ahmad, Naveen Suthar, H Haripriya, Pragya Kumar, Purnima Singh, Vijay Kumar, Sanjay Pandey, Madhusudan Prasad Singh

## Abstract

**Objective:** This study aimed to determine the national pooled prevalence of AUB, its sub-types, and dysmenorrhea among adolescent girls in India

**Methods:** We conducted a systematic review and meta-analysis following PRISMA guidelines, searching PubMed, Embase, and Web of Science from inception to December 4, 2024. Thirty-one observational studies involving adolescent girls aged 10-19 years with sample size ranging from 127 to 2000, conducted across India, were included. Due to high heterogeneity, pooled prevalence estimates were calculated using random-effects models.

**Results:** The pooled prevalence of AUB was 19% (95% CI: 15.0–22.0%). Pooled prevalence for the most common AUB sub-types noted are - oligomenorrhea (25%), metrorrhagia (24%), and menorrhagia (18%). Additionally, the pooled prevalence of dysmenorrhea was notably high, 53% (95% CI: 43–62%). Significant heterogeneity (I² > 90%) and regional disparities in prevalence were also observed in this review.

**Conclusion:** High prevalence of AUB (19 %) and its sub-types highlights the urgent need for the development of a standardised screening tool to be used among adolescent girls in schools and the community, and also the integration of screening programs in adolescent reproductive and sexual health clinics to combat long-term health burden

**What is already known on this subject?:** It has been previously established that abnormal uterine bleeding (AUB) and dysmenorrhea are serious gynecological issues in adolescent females worldwide, with studies indicating a variable prevalence. In India, several individual studies and regional reviews have shown the prevalence of various menstruation disorders, emphasizing concerns like oligomenorrhea related to PCOS and the high incidence of dysmenorrhea. These studies have consistently identified cultural stigma, lack of awareness, and diagnostic delays as significant impediments to effective care. The current literature is fragmented, exhibiting significant methodological variability, regional biases, and an absence of a unified national estimate, complicating the assessment of the overall burden of AUB and its specific subtypes among India’s adolescent population

**What this study adds?:** This systematic review and meta-analysis provide the inaugural comprehensive, national-level pooled prevalence estimates for AUB and its distinct subtypes solely among adolescent females in India. By combining data from 31 studies in different parts of India, we find that almost one-fifth (19%) of Indian teens have AUB. The most prevalent kinds are oligomenorrhea (25%) and metrorrhagia (24%), while more than half (53%) have dysmenorrhea. This study transcends narrative synthesis to provide substantial, quantitative evidence of pronounced regional differences and the considerable variation arising from diverse diagnosis criteria. Our findings highlight the essential public health necessity for the creation of standardized screening instruments and the incorporation of targeted interventions into school and community health initiatives to alleviate the enduring repercussions of these widespread illnesses.

## Introduction

Abnormal Uterine Bleeding (AUB) is characterized by uterine corpus bleeding that deviates from normal regularity, volume, frequency, or duration, occurring without pregnancy. (Munro et al., 2024) It is one of the most prevalent gynecological issues for teenage girls, and it has a big effect on their physical health, mental health, and schoolwork. (Elmaoğulları & Aycan, 2018) Persistent AUB can cause problems like anemia, depression, and long-term reproductive problems. It also hides other health problems, such as polycystic ovarian syndrome (PCOS), thyroid problems, and bleeding disorders (such von Willebrand disease).(Temuroğlu et al., 2025) In teenagers, the hypothalamic-pituitary-ovarian (HPO) axis is frequently underdeveloped, resulting in anovulatory cycles and erratic bleeding patterns throughout the earliest post-menarchal years. (Lemarchand-Béraud et al., 1982) Although temporary anomalies are prevalent, chronic abnormalities extending beyond 2-3 years of menarche necessitate clinical assessment.(Anthon et al., 2024) Even if this is true, diagnostic delays are common in India because of cultural stigmas, women’s lack of knowledge, and broken healthcare institutions. (Liu et al., 2007) For example, 90% of women with menorrhagia have bleeding issues (James, 2016), yet screening rates are still very low. (Jacobson et al., 2018) Likewise, oligomenorrhea and other sub-types of abnormal uterine bleeding (AUB) are significant symptoms associated with polycystic ovary syndrome (PCOS); one study has linked this symptom to an elevated risk of cardiovascular disease (CVD), coronary heart disease (CHD), and myocardial infarction (MI) in women.(Lo et al., 2023) Other types of abnormal uterine bleeding include hypermenorrhoea, hypomenorrhoea, and metrorrhagia. These conditions may go unnoticed, but they can have a big effect on young girls’ reproductive health in the long term. The PALM-COEIN method divides AUB into structural (polyp, adenomyosis, leiomyoma, malignancy/hyperplasia) and non-structural (coagulopathy, ovulatory dysfunction, endometrial, iatrogenic, not otherwise categorized) causes. (Munro et al., 2024) (Mikes et al., 2025) Dysmenorrhea is another prominent menstrual problem for teenage females. Even though therapy is available, it is still not recognized because of fear and cultural stigmas surrounding it. (Smith, 2018) Dysmenorrhea often accompanies abnormal uterine bleeding (AUB), compounding the difficulties, although both illnesses are typically underreported and poorly handled. To treat these illnesses in the Indian population, it is necessary to provide standardized protocols for early diagnosis and care. But this means that we need to figure out how to measure pooled estimates of AUB and its variants in India. For evidence-based healthcare planning, it’s important to know the real burden of AUB and its variants. There are a lot of different research on AUB and its sub-types in Indian teens, and they all use different methods and have regional biases. Consequently, we performed a comprehensive review and meta-analysis to ascertain the aggregated prevalence of Abnormal Uterine Bleeding (AUB) and its sub-types in teenage females in India. Also, looked at the pooled estimate dysmenorrhea among teenage girls in India.

## Methods

### Study Design

The goal of this systematic review was to find out the overall rate of Abnormal Uterine Bleeding (AUB) and its sub-types among teenage girls in India, following the 2020 standards from the Preferred Reporting Items for Systematic Reviews and Meta-Analyses (PRISMA). The protocol for this review was also recorded with the International Prospective Register of Systematic Reviews (PROSPERO). Number of registration: (CRD42024573458)

### Data Sources and Search Strategy

We made a thorough search plan to find all peer-reviewed papers for our systematic review and meta-analysis on three electronic databases: PubMed, Embase, and Web of Science. We looked for items that were written in English and published between inception to December 4, 2024. A systematic search of papers was conducted utilizing a combination of controlled vocabulary (MeSH/Emtree terms) and keywords associated with four principal concepts: (1) menstrual disorders (e.g., oligomenorrhea, dysmenorrhea, menorrhagia, PCOS, etc.), (2) adolescent populations (e.g., “adolescent,”“school-going girls,” etc.), (3) geographical focus (India), and (4) epidemiological terms (e.g., prevalence, burden). (See Table 1) The search turned up 514 records from PubMed, 326 from Embase, and 183 from Web of Science. All searches were run from inception to December 4, 2024, across three databases, and all identified records were included for abstract screening. Before the screening process began, the retrieved records were brought into Ryaan Software to get rid of any duplicates.

**Table 1:**
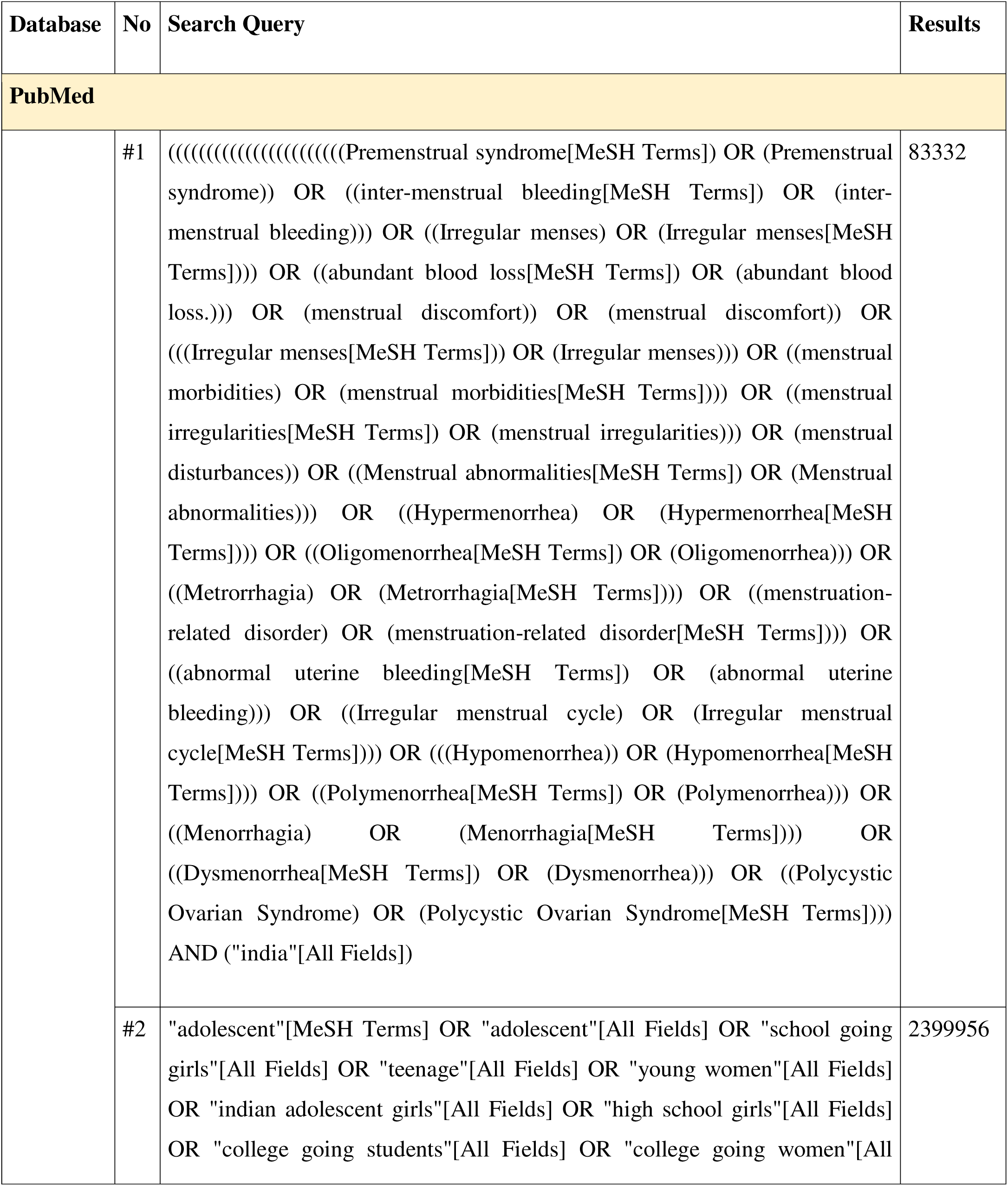
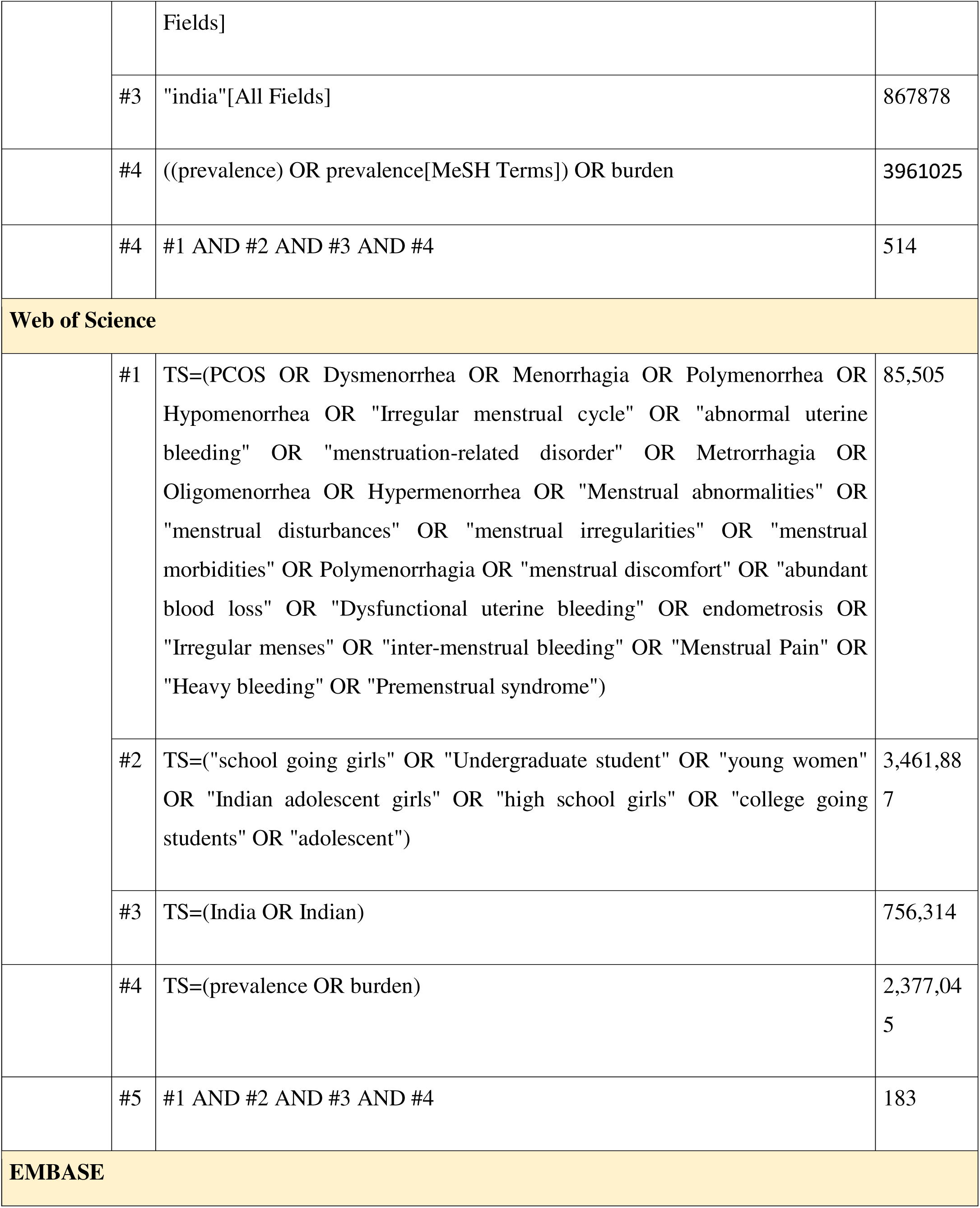
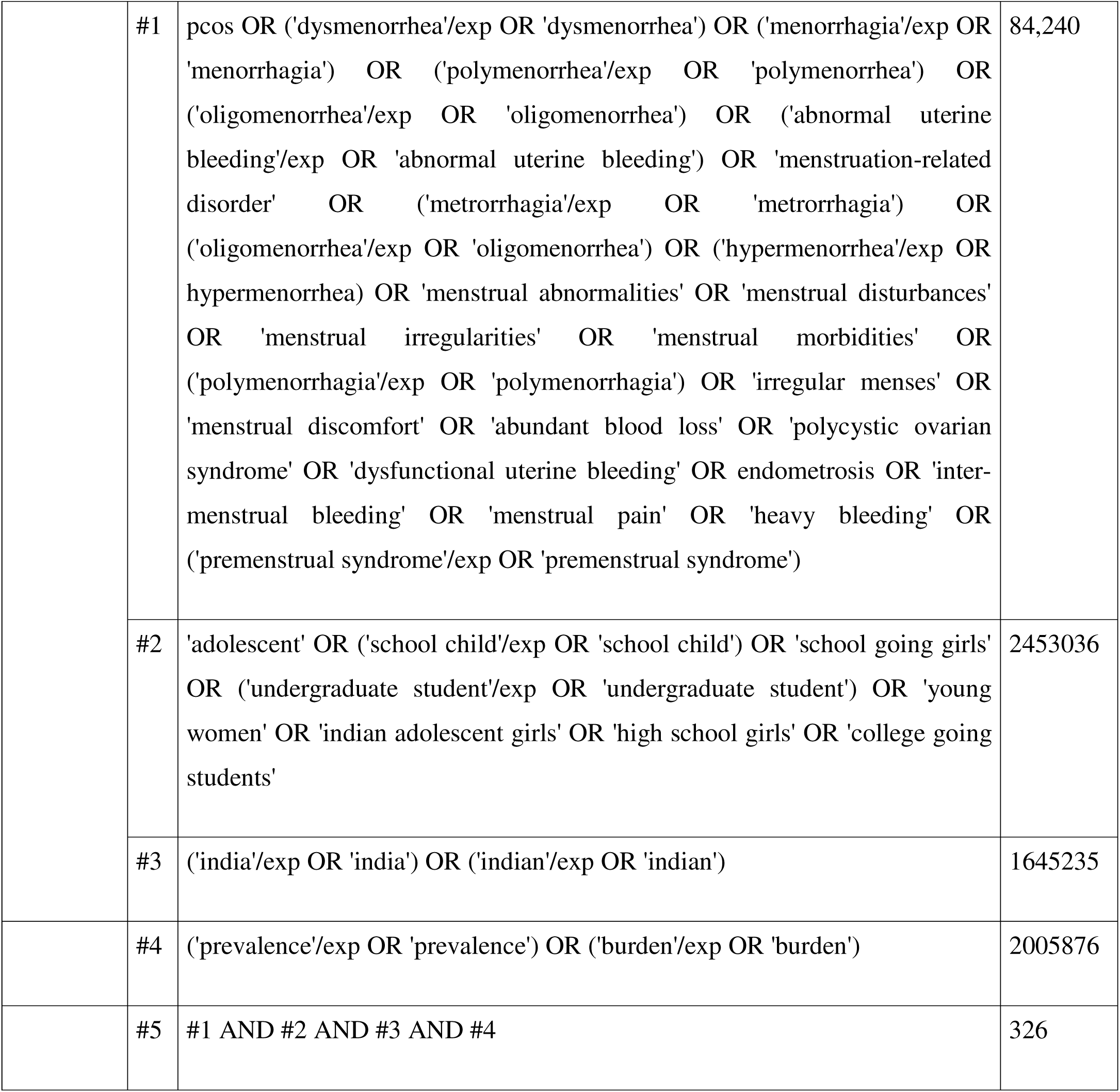
Search strategy for the review.

| Database | No | Search Query | Results |
| --- | --- | --- | --- |
| **PubMed** |  |  |  |
|  | #1 | ((((((((((((((((((((Premenstrual syndrome[MeSH Terms]) OR (Premenstrual syndrome)) OR ((inter-menstrual bleeding[MeSH Terms]) OR (inter-menstrual bleeding))) OR ((Irregular menses) OR (Irregular menses[MeSH Terms]))) OR ((abundant blood loss[MeSH Terms]) OR (abundant blood loss.))) OR (menstrual discomfort)) OR (menstrual discomfort)) OR (((Irregular menses[MeSH Terms]) OR (Irregular menses))) OR ((menstrual morbidities) OR (menstrual morbidities[MeSH Terms]))) OR ((menstrual irregularities[MeSH Terms]) OR (menstrual irregularities))) OR (menstrual disturbances)) OR ((Menstrual abnormalities[MeSH Terms]) OR (Menstrual abnormalities))) OR ((Hypermenorrhea) OR (Hypermenorrhea[MeSH Terms]))) OR ((Oligomenorrhea[MeSH Terms]) OR (Oligomenorrhea))) OR ((Metrorrhagia) OR (Metrorrhagia[MeSH Terms]))) OR ((menstruation-related disorder) OR (menstruation-related disorder[MeSH Terms]))) OR ((abnormal uterine bleeding[MeSH Terms]) OR (abnormal uterine bleeding))) OR ((Irregular menstrual cycle) OR (Irregular menstrual cycle[MeSH Terms]))) OR (((Hypomenorrhea) OR (Hypomenorrhea[MeSH Terms]))) OR ((Polymenorrhea[MeSH Terms]) OR (Polymenorrhea))) OR ((Menorrhagia) OR (Menorrhagia[MeSH Terms])) OR ((Dysmenorrhea[MeSH Terms]) OR (Dysmenorrhea))) OR ((Polycystic Ovarian Syndrome) OR (Polycystic Ovarian Syndrome[MeSH Terms]))) AND ("india"[All Fields]) | 83332 |
|  | #2 | "adolescent"[MeSH Terms] OR "adolescent"[All Fields] OR "school going girls"[All Fields] OR "teenage"[All Fields] OR "young women"[All Fields] OR "indian adolescent girls"[All Fields] OR "high school girls"[All Fields] OR "college going students"[All Fields] OR "college going women"[All | 2399956 |
|  |  | Fields] |  |
|  | #3 | "india"[All Fields] | 867878 |
|  | #4 | ((prevalence) OR prevalence[MeSH Terms]) OR burden | 3961025 |
|  | #4 | #1 AND #2 AND #3 AND #4 | 514 |
| Web of Science |  |  |  |
|  | #1 | TS=(PCOS OR Dysmenorrhea OR Menorrhagia OR Polymenorrhea OR Hypomenorrhea OR "Irregular menstrual cycle" OR "abnormal uterine bleeding" OR "menstruation-related disorder" OR Metrorrhagia OR Oligomenorrhea OR Hypermenorrhea OR "Menstrual abnormalities" OR "menstrual disturbances" OR "menstrual irregularities" OR "menstrual morbidities" OR Polymenorrhagia OR "menstrual discomfort" OR "abundant blood loss" OR "Dysfunctional uterine bleeding" OR endometriosis OR "Irregular menses" OR "inter-menstrual bleeding" OR "Menstrual Pain" OR "Heavy bleeding" OR "Premenstrual syndrome") | 85,505 |
|  | #2 | TS=("school going girls" OR "Undergraduate student" OR "young women" OR "Indian adolescent girls" OR "high school girls" OR "college going students" OR "adolescent") | 3,461,887 |
|  | #3 | TS=(India OR Indian) | 756,314 |
|  | #4 | TS=(prevalence OR burden) | 2,377,045 |
|  | #5 | #1 AND #2 AND #3 AND #4 | 183 |

EMBASE
|  |  |  |
| --- | --- | --- |
| #1 | pcos OR ('dysmenorrhea'/exp OR 'dysmenorrhea') OR ('menorrhagia'/exp OR 'menorrhagia') OR ('polymenorrhea'/exp OR 'polymenorrhea') OR ('oligomenorrhea'/exp OR 'oligomenorrhea') OR ('abnormal uterine bleeding'/exp OR 'abnormal uterine bleeding') OR 'menstruation-related disorder' OR ('metrorrhagia'/exp OR 'metrorrhagia') OR ('oligomenorrhea'/exp OR 'oligomenorrhea') OR ('hypermenorrhea'/exp OR hypermenorrhea) OR 'menstrual abnormalities' OR 'menstrual disturbances' OR 'menstrual irregularities' OR 'menstrual morbidities' OR ('polymenorrhagia'/exp OR 'polymenorrhagia') OR 'irregular menses' OR 'menstrual discomfort' OR 'abundant blood loss' OR 'polycystic ovarian syndrome' OR 'dysfunctional uterine bleeding' OR endometrosis OR 'intermenstrual bleeding' OR 'menstrual pain' OR 'heavy bleeding' OR ('premenstrual syndrome'/exp OR 'premenstrual syndrome') | 84,240 |
| #2 | 'adolescent' OR ('school child'/exp OR 'school child') OR 'school going girls' OR ('undergraduate student'/exp OR 'undergraduate student') OR 'young women' OR 'indian adolescent girls' OR 'high school girls' OR 'college going students' | 2453036 |
| #3 | ('india'/exp OR 'india') OR ('indian'/exp OR 'indian') | 1645235 |
| #4 | ('prevalence'/exp OR 'prevalence') OR ('burden'/exp OR 'burden') | 2005876 |
| #5 | #1 AND #2 AND #3 AND #4 | 326 |

### Eligibility criteria

We chose studies based on certain criteria that had already been set. We incorporated observational studies (cross-sectional, cohort, and case-control) and randomized controlled trials with inclusion of baseline prevalence data on outcomes concerning teenage females aged 10-19 years in India. We concentrated on studies that included data regarding outcomes pertinent to this research, including the prevalence of abnormal uterine bleeding and its various sub-types (dysmenorrhea, oligomenorrhea, polymenorrhea, etc.). We didn’t include studies that focused on teens who were pregnant . This review were not done on populations that weren’t Indian, or didn’t have extractable prevalence data that was relevant to our study outcomes. We only looked at publications that were in English. Also, case reports, review articles, and publications that weren’t peer-reviewed were left out. We only chose papers that were published from the inception upto December 4, 2024.

#### Data Extraction Using Rayyan Software

Two independent reviewers looked at the publications using Rayyan software. The reviewers systematically evaluated the titles and abstracts against the eligibility criteria to see methodological rigor. The discrepancies in article selection and data extraction were addressed through consultation with a third reviewer. We then got the full texts of papers that might be suitable and uploaded them to Rayyan software. The recovered articles were meticulously re-evaluated to ascertain their eligibility, and pertinent data were meticulously collected. We utilized a standardized data extraction form to gather information about the study’s characteristics (author, year, state, study design, sample size, and study setting), population (sample size and age distribution), and prevalence estimates for each type of AUB (operational definitions used for each type, etc.).

Rayyan’s capabilities, such as blind review mode, tagging, and collaborative tools, made it easy to screen massive sets of results quickly. All studies that were not included were written down with explanations for why they were not included to keep things clear. The software’s export capability made it possible to make flowcharts that follow PRISMA standards to show how the research selection process worked.

### Data Analysis

The statistical analysis utilized the “meta” package in R Studio, establishing statistical significance at a P value < 0.05. We used a random-effects meta-analysis model with inverse-variance weighting to account for differences between studies in order to estimate the overall prevalence, type-specific prevalence (oligomenorrhea, menorrhagia, polymenorrhea, hypomenorrhea, and metrorrhagia), and a secondary analysis of dysmenorrhea. We used I² statistics and Cochran’s Q test to measure heterogeneity. An I² value of 75% or higher meant that there was a lot of heterogeneity. To explore possible sources of variation, subgroup analyses were performed according to geographic location in India (North, South, East, West) and sample size (<500 vs. ≥500 participants). Funnel plots and Egger’s regression test were used to look for publication bias. A p-value of less than 0.05 means that there is significant publication bias.

## Result

There were 1,023 records found in databases and registrations. After removing 166 duplicates, there were 857 records left for screening. After the first screening, 742 data were excluded, and 115 full-text reports were requested for retrieval. There were 28 reports that couldn’t be found, thus, there were only 87 records left to check for eligibility. (Figure 1) After evaluation, 57 papers were removed, mostly because they didn’t meet the selection criteria (n=46), had the improper findings (n=9), or were a scoping review (n=1). In the end, 31 studies satisfied the criteria for inclusion and were included for systematic review.

**Figure 1:**
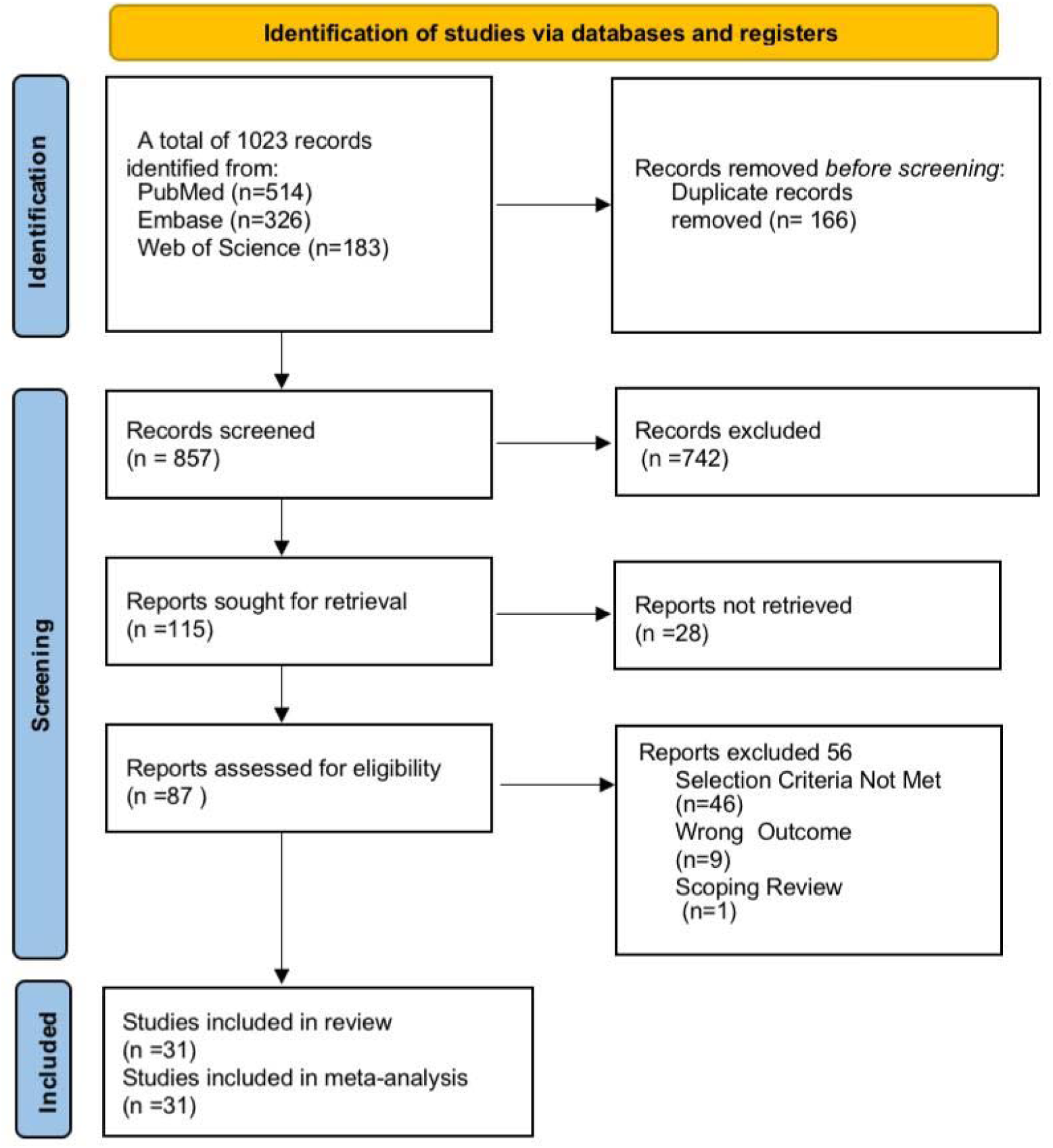
Search stategy using various databases.

### Characteristics of included studies

The systematic review and meta-analysis comprised 31 studies conducted in several regions of India, involving a total of 15,678 adolescent girls aged 10 to 19 years. The majority of the studies were cross-sectional (n=27), while the others were cohort (n=2), prospective analytic (n=1), and case-control (n=1) investigations. The size of samples ranged from 127 to 2,000. The studies were conducted in North India (n=10), South India (n=10), West India (n=6) and East India (n=5). (Table 2)

**Table 2:** Characteristics and quality assessment of studies.

| **Sr No.** | **Name of the study author** | **Year of Publication** | **State** | **Type of study design** | **Sample size** | **Age Group (in yrs)** | **Abnormal Uterine Bleeding in Participants** |
| --- | --- | --- | --- | --- | --- | --- | --- |
| 1. | (Rani et al., 2016) | 2015 | Chandigarh | Cross-sectional | 300 | 10-19 | Dysmenorrhoea-184<br>Oligomenorrhea-94<br>Polymenorrhoea-19 |
| 2. | (Rathod et al., 2016) | 2016 | Maharashtra | Prospective and analytic study | 655 | 11-19 | Dysmenorrhoea-213,<br>Oligomenorrhea-192,<br>Menorrhagia-97 |
| 3. | (Pegu et al., 2018) | 2018 | Andaman and Nicobar Islands | Descriptive analysis | 824 | 10-19 | Dysmenorrhoea-99,<br>Oligomenorrhea-92,<br>Polymenorrhoea-75,<br>Menorrhagia-138 |
| 4. | (Dambhare et al., 2012) | 2012 | Maharashtra | Cross-sectional | 561 | 10-19 | Dysmenorrhoea-315,<br>Oligomenorrhea-124,<br>Polymenorrhoea-47,<br>Hypermenorrhoea-7,<br>Hypomenorrhoea-9, |
| 5. | (Tarannum et al., 2021) | 2021 | Uttar Pradesh | Cross-sectional | 422 | 10-19 | Oligomenorrhea-119,<br>Polymenorrhoea-63,<br>Hypermenorrhoea-109, |
|  |  |  |  |  |  |  | Hypomenorrhoea-16,<br>Menorrhagia-172,<br>Metrorrhagia-185 |
| 6. | (Wadgave et al., 2014) | 2014 | Maharashtra | Cross-sectional | 400 | 13–19 | Dysmenorrhea-180,<br>Oligomenorrhoea-61,<br>Polymenorrhoea-33,<br>Menorrhagia-55 |
| 7. | (I. Sharma et al., 2015) | 2015 | New Delhi | Cohort | 280 | 10-19 | Dysmenorrhea-20,<br>Oligomenorrhoea-30,<br>Menorrhagia-82,<br>Metrorrhagia-55 |
| 8. | (Shringarpure et al., 2023) | 2023 | Gujarat | Cross-sectional | 308 | 10-19 | Dysmenorrhea-118,<br>Oligomenorrhoea-43,<br>Menorrhagia-26, |
| 9. | (K. Verma & Baniya, 2022) | 2022 | Rajasthan | Cross-sectional | 492 | 13-19 | Dysmenorrhea-318 |
| 10. | (Madaan et al., 2014) | 2010 | Delhi | Cross-sectional | 316 | 11-19 | Dysmenorrhea-118 |
| 11. | (M. Singh et al., 2019) | 2019 | Delhi | Cross-sectional | 210 | 10-19 | Dysmenorrhea-138,<br>Oligomenorrhea-11,<br>Polymenorrhoea-11,<br>Hypermenorrhoea-53<br>Hypomenorrhoea-9,<br>Menorrhagia-53 |
| 12. | (Agarwal et al., 2024) | 2024 | Bihar, India | Cross-sectional | 2000 | 10 - 19 | Dysmenorrhea-315,<br>Oligomenorrhea-215,<br>Polymenorrhoea-28<br>Hypermenorrhoea-53,<br>Hypomenorrhoea-43<br>Menorrhagia-304<br>Metrorrhagia-184 |
| 13. | (S et al., 2020) | 2014 | Karnataka | Cross-sectional | 233 | 12 - 17 years | Dysmenorrhea-146<br>Hypomenorrhoea-146<br>Oligomenorrhoea-77<br>Hypermenorrhoea-7 |
| 14. | (P. Singh et al., 2016) | 2016 | Uttar Pradesh | Cross-sectional | 260 | Age not mentioned, but includes adolescent girls | Dysmenorrhea-123 |
| 15. | (P. Sharma et al., 2008) | 2008 | Delhi | Cross-sectional | 198 | 13-19 years | Dysmenorrhea-133<br>Hypomenorrhoea-9<br>Menorrhagia-27<br>Hypermenorrhoea-30<br>Oligomenorrhoea-63 |
| 16. | (Mathiyalagen et al., 2017) | 2017 | Puducherry, India | Cross-sectional | 242 | 12-18 | Dysmenorrhea-199<br>Hypomenorrhoea-5<br>Menorrhagia-55<br>Hypermenorrhoea-51 |
| 17. | (Mukherjee et al., 2014) | 2014 | West Bengal | Cross-sectional | 159 | 15-19 | Dysmenorrhea-90,<br>Oligomenorrhea-36<br>Hypermenorrhoea-50 |
| 18. | (Negi et al., 2018) | 2018 | Uttarakhand | Cross-sectional | 470 | 13-19 | Dysmenorrhea-295<br>Menorrhagia-45<br>Oligomenorrhea-135 |
| 19. | (Yaliwal et al., 2020) | 2020 | Karnataka | Cross-sectional | 1016 | 10-19 | Dysmenorrhea-630<br>Hypomenorrhoea-118<br>Hypermenorrhoea-91<br>Oligomenorrhea- 221<br>Menorrhagia-124 |
| 20. | (Nidhi et al., 2011) | 2011 | Andhra Pradesh | Cohort | 460 | 15-18 | Oligomenorrhea-72 |
| 21. | (Ravi et al., 2017) | 2017 | Tamil Nadu | Cross-sectional | 350 | 10 -19 | Dysmenorrhea-254<br>Menorrhagia-160 |
| 22. | (Juyal et al., 2023) | 2014 | Uttarakhand | Cross-sectional | 453 | 15-16 | Dysmenorrhea-294<br>Oligomenorrhea-16<br>Hypomenorrhoea-48 |
| 23. | (Vani K et al., 2013) | 2013 | Pondicherry | Cross-sectional | 853 | 13-19 | Dysmenorrhea-621<br>Oligomenorrhea-149 |
| 24. | (Sanyal & Ray, 2008) | 2008 | West Bengal | Cross-sectional | 220 | 14-18 | Dysmenorrhea-166<br>Oligomenorrhea-209<br>Hypermenorrhoea-84 |
| 25. | (Omidvar et al., 2015) | 2018 | Karnatak | Cross-sectional | 536 | 10-19 | Dysmenorrhea-356 |
|  |  |  |  |  |  |  | Hypermenorrhoea-64<br>Oligomenorrhoea-53<br>Polymenorrhoea-79<br>Menorrhagia-159 |
| 26. | (Rajiwade et al., 2018) | 2017 | Puducherry | Case-control | 144 | 10–19 | Dysmenorrhea-26 |
| 27. | (Ray et al., 2010) | 2010 | West Bengal | Cross-sectional | 715 | 10 - 19 | Dysmenorrhea-218<br>Oligomenorrhea-639<br>Menorrhagia-131 |
| 28. | (Thakre et al., 2012) | 2012 | Maharashtra | Cross-sectional | 387 | 12 -17 | Dysmenorrhea-236<br>Hypomenorrhoea-4<br>Menorrhagia-35<br>Oligomenorrhea-58 |
| 29. | (Sharanya, 2014) | 2014 | Tamil Nadu | Cross-sectional | 130 | 13 -19 | Dysmenorrhea-79<br>Polymenorrhoea-27<br>Menorrhagia-41 |
| 30. | (Parikh & Nagar, 2022) | 2022 | Gujarat | Cross-sectional | 127 | 18 (0.70) | Dysmenorrhea-82<br>Oligomenorrhoea-25 |
|  |  |  |  |  |  |  | Hypomenorrhoea-2<br>Hypermenorrhoea-6<br>Menorrhagia-14 |
| 31 | (Khan et al., 2024) | 2024 | Uttar Pradesh | Cross-sectional | 640 | 10 - 19 | Dysmenorrhea-472<br>Oligomenorrhoea-71<br>Polymenorrhoea-65<br>Hypomenorrhoea-179 |

**Table 3:** Quality assessment using Joanna Briggs Institute (JBI) checklist.

| Sr No. | Name of the Author & Year of Publication | 1.Were the criteria for inclusion in the sample clearly defined? | 2.Were the study subjects and the setting described in detail? | 3.Was the exposure measured in a valid and reliable way? | 4.Were objective, standard criteria used for measurement of the condition? | 5. Were confounding factors identified? | 6.Were strategies to deal with confounding factors stated? | 7.Were the outcomes measured in a valid and reliable way? | 8.Was appropriate statistical analysis used? | T S |
| --- | --- | --- | --- | --- | --- | --- | --- | --- | --- | --- |
| 1. | (Rani et al., 2016) | 1 | 1 | 1 | 1 | 0 | 0 | 1 | 1 | 6 |
| 2. | (Rathod et al., 2016) | 1 | 1 | 1 | 1 | 0 | 0 | 1 | 1 | 6 |
| 3. | (Pegu et al., | 1 | 1 | 0 | 0 | 0 | 0 | 0 | 1 | 3 |
|  | 2018) |  |  |  |  |  |  |  |  |  |
| **4.** | (Dambh<br>are et<br>al.,<br>2012) | **1** | **1** | **1** | **0** | **0** | **0** | **1** | **1** | **5** |
| **5.** | (Tarannu<br>m et al.,<br>2021) | **1** | **1** | **1** | **0** | **1** | **1** | **1** | **1** | **7** |
| **6.** | (Wadgav<br>e et al.,<br>2014) | **1** | **1** | **1** | **1** | **0** | **0** | **1** | **1** | **6** |
| **7.** | (I.<br>Sharma<br>et al.,<br>2015) | **1** | **1** | **1** | **1** | **0** | **0** | **1** | **1** | **6** |
| **8.** | (Shringar<br>pure et<br>al.,<br>2023) | **1** | **1** | **0** | **1** | **0** | **0** | **0** | **1** | **4** |
| **9.** | (K.<br>Verma &<br>Baniya,<br>2022) | **1** | **1** | **1** | **1** | **0** | **0** | **1** | **1** | **6** |
| **10.** | (Madaan<br>et al.,<br>2014) | **1** | **1** | **0** | **0** | **1** | **0** | **0** | **1** | **3** |
| **11.** | (M.<br>Singh et<br>al.,<br>2019) | **1** | **1** | **1** | **0** | **1** | **0** | **0** | **1** | **5** |
| **12.** | (Agarwal<br>et al., | **1** | **1** | **1** | **1** | **1** | **0** | **1** | **1** | **7** |
|  | 2024) |  |  |  |  |  |  |  |  |  |
| **13.** | (S et al., 2020) | **1** | **1** | **1** | **0** | **1** | **0** | **1** | **1** | **6** |
| **14.** | (P. Singh et al., 2016) | **1** | **1** | **1** | **1** | **0** | **0** | **1** | **1** | **6** |
| **15.** | (P. Sharma et al., 2008) | **1** | **1** | **1** | **0** | **0** | **0** | **1** | **1** | **5** |
| **16.** | (Mathiyalagen et al., 2017) | **1** | **1** | **1** | **0** | **0** | **0** | **1** | **1** | **5** |
| **17.** | (Mukherjee et al., 2014) | **1** | **1** | **1** | **0** | **1** | **1** | **0** | **1** | **6** |
| **18.** | (Negi et al., 2018) | **1** | **1** | **0** | **0** | **1** | **0** | **0** | **1** | **4** |
| **19.** | (Yaliwal et al., 2020) | **1** | **1** | **0** | **0** | **1** | **0** | **1** | **1** | **5** |
| **20.** | (Nidhi et al., 2011) | **1** | **1** | **1** | **1** | **0** | **0** | **1** | **1** | **6** |
| **21.** | (Ravi et al., 2017) | **1** | **1** | **1** | **1** | **0** | **0** | **1** | **1** | **6** |
| **22.** | (Juyal et al., 2023) | **1** | **1** | **1** | **1** | **0** | **0** | **1** | **1** | **6** |
| **23.** | (Vani K<br>et al.,<br>2013) | **1** | **1** | **0** | **1** | **1** | **0** | **0** | **1** | **5** |
| **24.** | (Sanyal<br>& Ray,<br>2008) | **1** | **1** | **1** | **0** | **1** | **1** | **0** | **1** | **6** |
| **25.** | (Omidva<br>r et al.,<br>2015) | **1** | **1** | **1** | **0** | **0** | **0** | **0** | **1** | **4** |
| **26.** | Rajiwad<br>e et al.,<br>2018) | **1** | **1** | **1** | **1** | **0** | **0** | **1** | **1** | **6** |
| **27.** | (Ray et<br>al.,<br>2010) | **1** | **1** | **0** | **0** | **1** | **1** | **0** | **1** | **5** |
| **28.** | (Thakre<br>et al.,<br>2012) | **1** | **1** | **0** | **0** | **1** | **1** | **0** | **1** | **5** |
| **29.** | (Sharany<br>a, 2014) | **1** | **1** | **1** | **0** | **0** | **0** | **0** | **1** | **4** |
| **30.** | (Parikh<br>& Nagar,<br>2022) | **1** | **1** | **0** | **0** | **0** | **0** | **0** | **1** | **3** |
| **31.** | (Khan et<br>al.,<br>2024) | **1** | **1** | **0** | **0** | **0** | **0** | **0** | **1** | **3** |

The quality assessment of the included 31 studies was done using the Joanna Briggs Institute (JBI) checklist for observational studies. Each study was scored across eight domains, a) clear inclusion criteria b) detailed description of subjects/settings, c) valid/reliable measurement of exposure and outcome measurement, d) use of standard diagnostic criteria for measurement of exposure and outcome measurement, e) identification of confounders, f) strategies to address confounding, g) valid and reliable way of outcome measurement h) appropriate statistical analysis, with items scored as 1 (yes) or 0 (no); total scores ranged from 3 to 7. Risk-of-bias Quality of assessments for risk of bias were performed independently by three authors, and help of fourth author was taken to resolve any conflicts arose.

### Pooled prevalence estimates of Abnormal Uterine Bleeding (AUB) among adolescents in India

The meta-analysis revealed a pooled prevalence of abnormal uterine bleeding (AUB) of 19% (95% CI: 15.4%–22.1%), indicating a substantial burden of this condition among the studied populations. The random-effects model was employed due to significant heterogeneity across studies (I² = 99.48%, p < 0.001), reflecting variability related to diagnostic criteria, regional influence, and sample size differences. The high Tau² value (0.0192, SE = 0.0076) further highlights the substantial between-studies variance which indicates cautious interpretation of the pooled estimate. Despite the high heterogeneity, the Z -score (10.9, p < 0.001) indicates statistical significance of the overall prevalence. (**Figure 2)**

**Figure 2:**
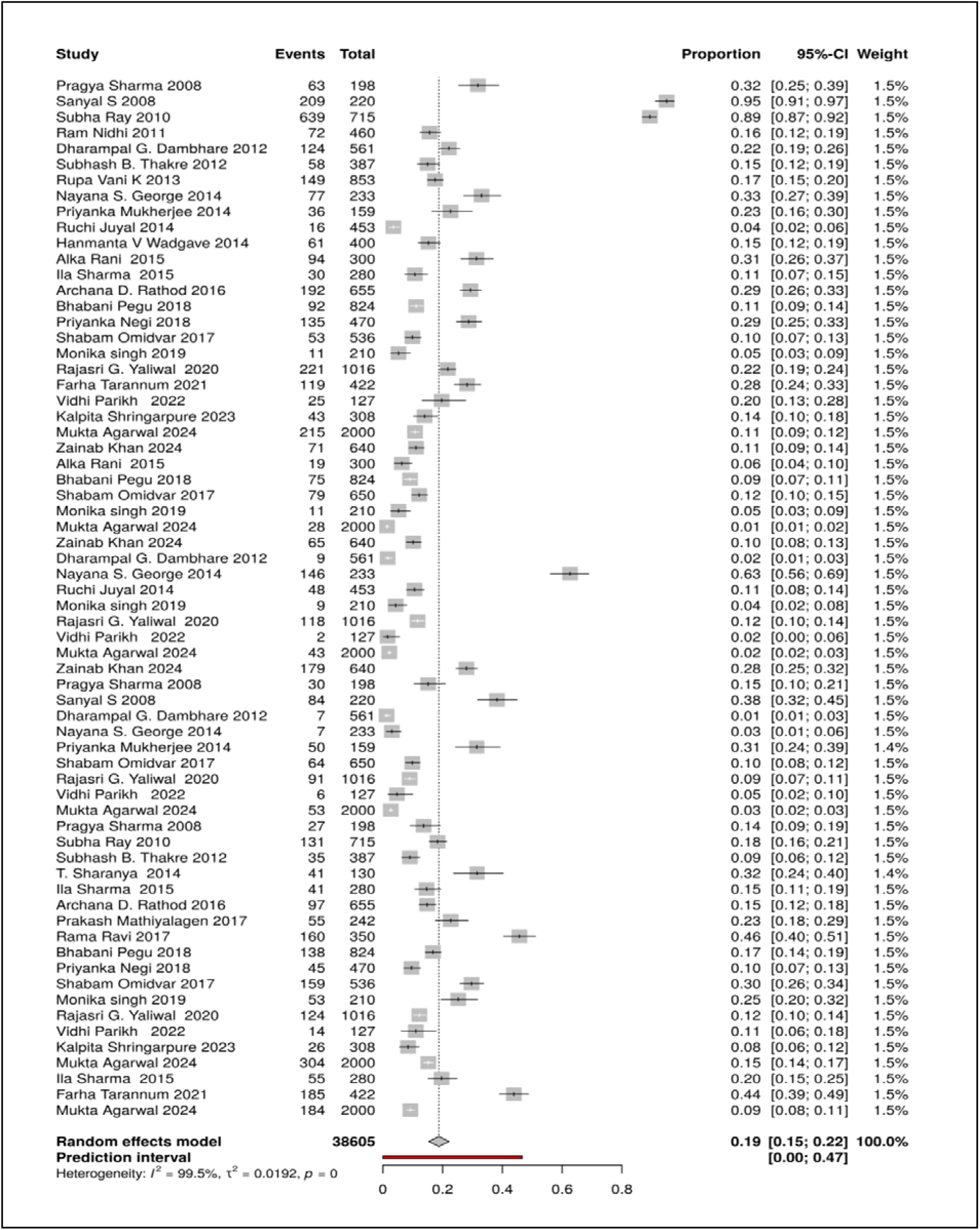
Forest plot showing pooled prevalence of Abnormal Uterine Bleeding (AUB) among Adolescent girls in India.

Visual representation through forest plots demonstrated the distribution of individual study estimates, most of which fell within our calculated confidence interval. **(Figure 3)** Funnel plot analysis revealed some asymmetry (Kendall’s τ = 0.346, p < 0.001; Egger’s Z = 5.04, p < 0.001), suggesting possible small-study effects. However, the remarkably high fail-safe N (196,616) provides strong evidence that our pooled estimate remains robust against potential publication bias.

**Figure 3:**
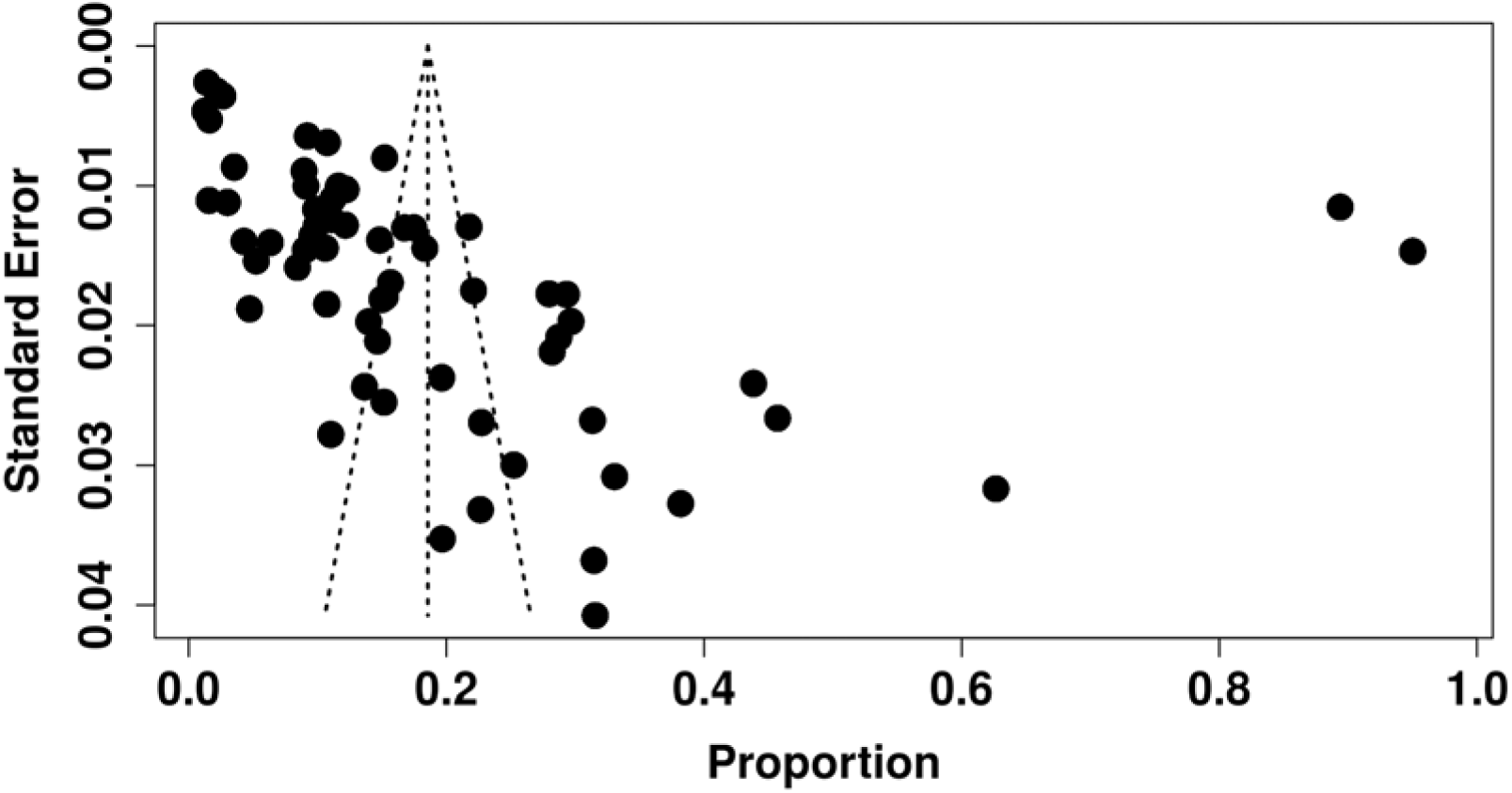
Funnel plot for the assessment of publication bias in the pooled prevalence of Abnormal Uterine Bleeding (AUB).

### Pooled prevalence estimates for several types of Abnormal Uterine Bleeding (AUB) among adolescent girls in India

Our subgroup analysis of the AUB sub-type revealed significant variance in the pooled prevalence among Indian adolescents across different states. The study revealed the greatest pooled prevalence for Oligomenorrhea at 25% (95% CI: 16-34%), with significant heterogeneity (I²=99.7%, τ²=0.0505, p=0). One study (Sanyal & Ray, 2008) found that the highest prevalence of oligomenorrhea was 95%. The lowest rate of oligomenorrhea recorded in the study Juyal (4%) (Juyal et al., 2023).

Following that, the high pooled prevalence for metrorrhagia was 24% (95% CI: 4-44%; range: 9-44%), with a high level of heterogeneity (I²=99%, τ²=0.0312, p<0.0001). The study by Tarannum et al. (44%) had the greatest prevalence of metrorrhagia (Tarannum et al., 2021), while the study by Agarwal et al. noted the lowest prevalence (9%) (Agarwal et al., 2024). The overall prevalence for Menorrhagia was 18% (95% CI: 14-23%) with significantheterogeneity (I²=94.8%, τ²=0.0093, p<0.0001), ranging from (Shringarpure et al., 2023) (8%) to Rama Ravi 2017 (46%) (Ravi et al., 2017). In the same way, Hypermenorrhea estimates were pooled at 15% (95% CI: 1-30%; range: 2-63%) with strong heterogeneity (I²=98.9%, τ²=0.0431, p<0.0001), with the lowest being 2% (Dambhare et al., 2012) and the highest being 63% (S et al., 2020). Hypomenorrhea, another significant sub-type of AUB, with a pooled prevalence of 12% (95% CI: 4-21%), characterized by considerable heterogeneity (I²=97.2%, τ²=0.0164, p<0.0001). The prevalence varies between 1% (Dambhare et al., 2012) to 38% (Sanyal & Ray, 2008).

Additionally, another sub-type, polymenorrhea, exhibited the lowest burden at 7% (95% CI: 4-11%; range: 1-12%) in subgroup analysis, although shown significant heterogeneity (I²=97%, τ²=0.0015, p<0.0001), with prevalence rates ranging from 1% (Agarwal et al., 2024) to 12% (Omidvar et al., 2015). The considerable subgroup differences (χ²=24.11, p=0.0002) corroborated geographical variances, notably in Eastern India, where a research by (Sanyal & Ray, 2008) indicated a 95% prevalence of oligomenorrhoea, marking it as an outlier and suggesting potential variations due to regional, methodological, and diagnostic factors. (Figure 4) Subgroup analysis has shown statistically significant, substantial heterogeneity (I² = 99.5%, p < 0.001), subgroup analyses were done to look for possible causes of the differences. The sub-group analysis indicated that no significant differences were found between studies with ≤500 participants and those with >500 participants (χ² = 0.36, p = 0.549). Moreover to it, notable geographical disparities were identified (χ² = 8.09, p = 0.0442), affirming that the frequency of AUB differs throughout India. Furthermore, the greatest diversity observed in the Eastern area of India suggests that additional region-specific factors (e.g., healthcare access, operational definition) affect prevalence.

**Figure 4:**
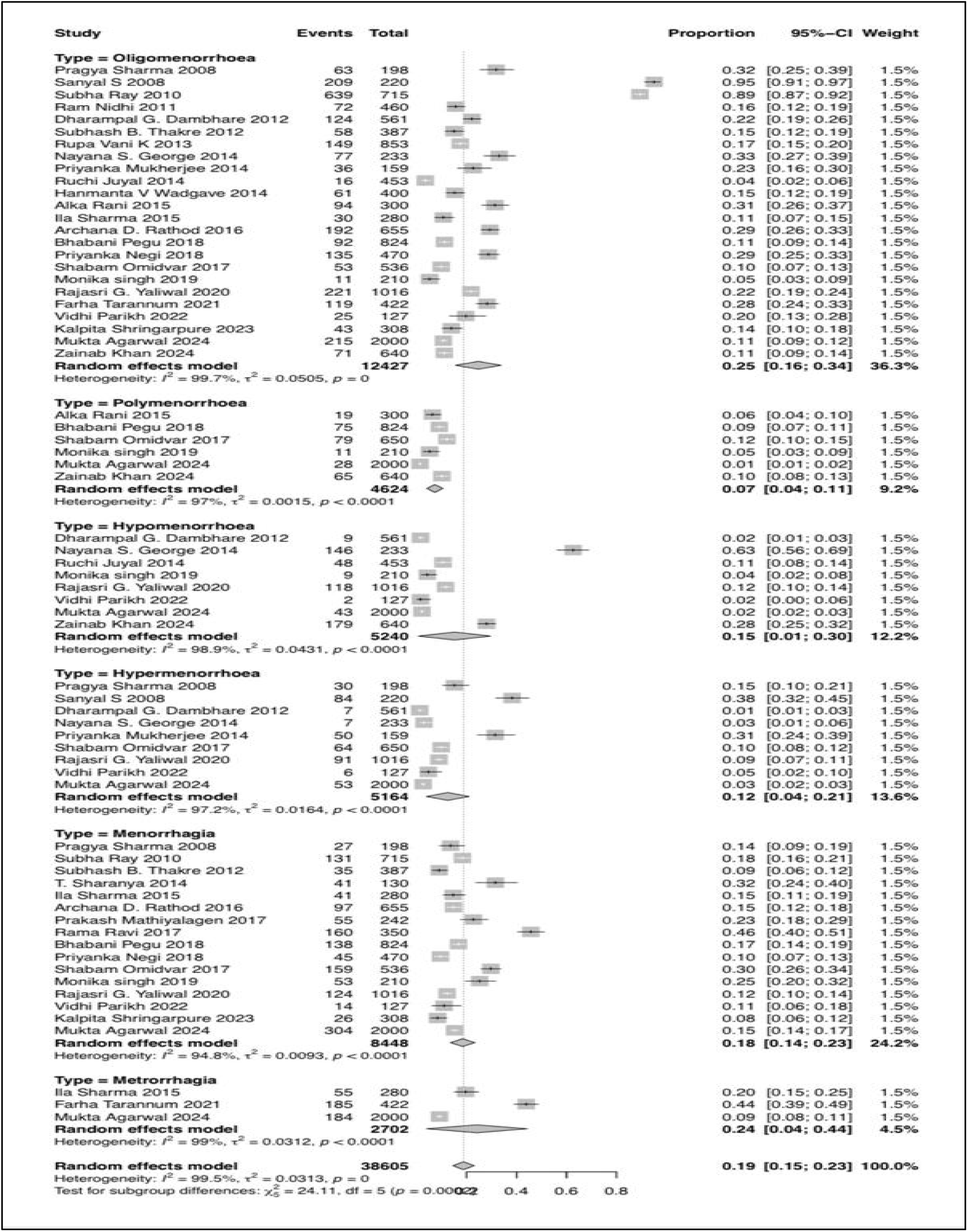
Subgroup analysis and Pooled prevalence for various type of Abnormal Uterine Bleeding among Adolescent girls in India.

**Figure 5:**
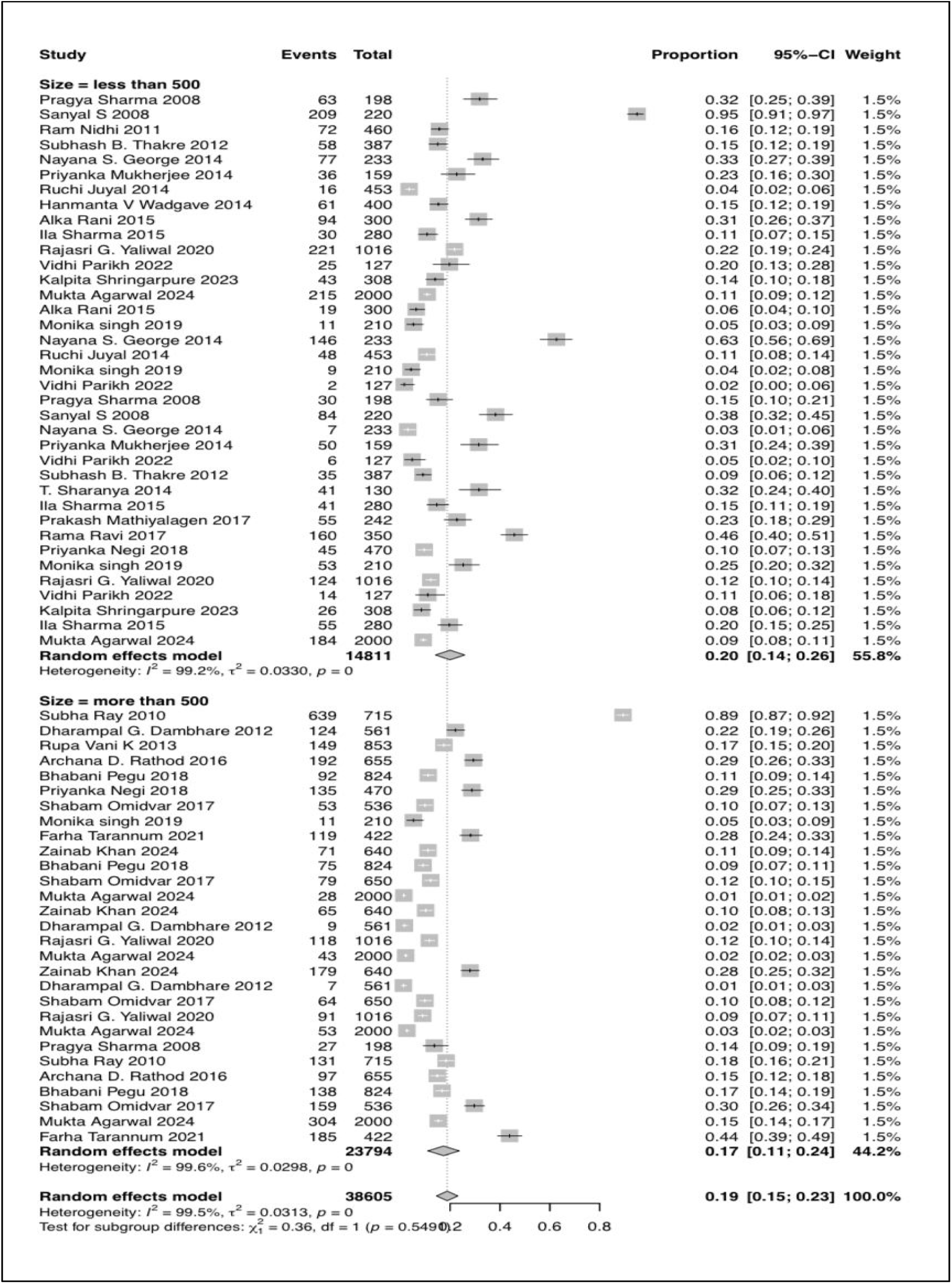
Sub-group –analysis by sample size.

**Figure 6:**
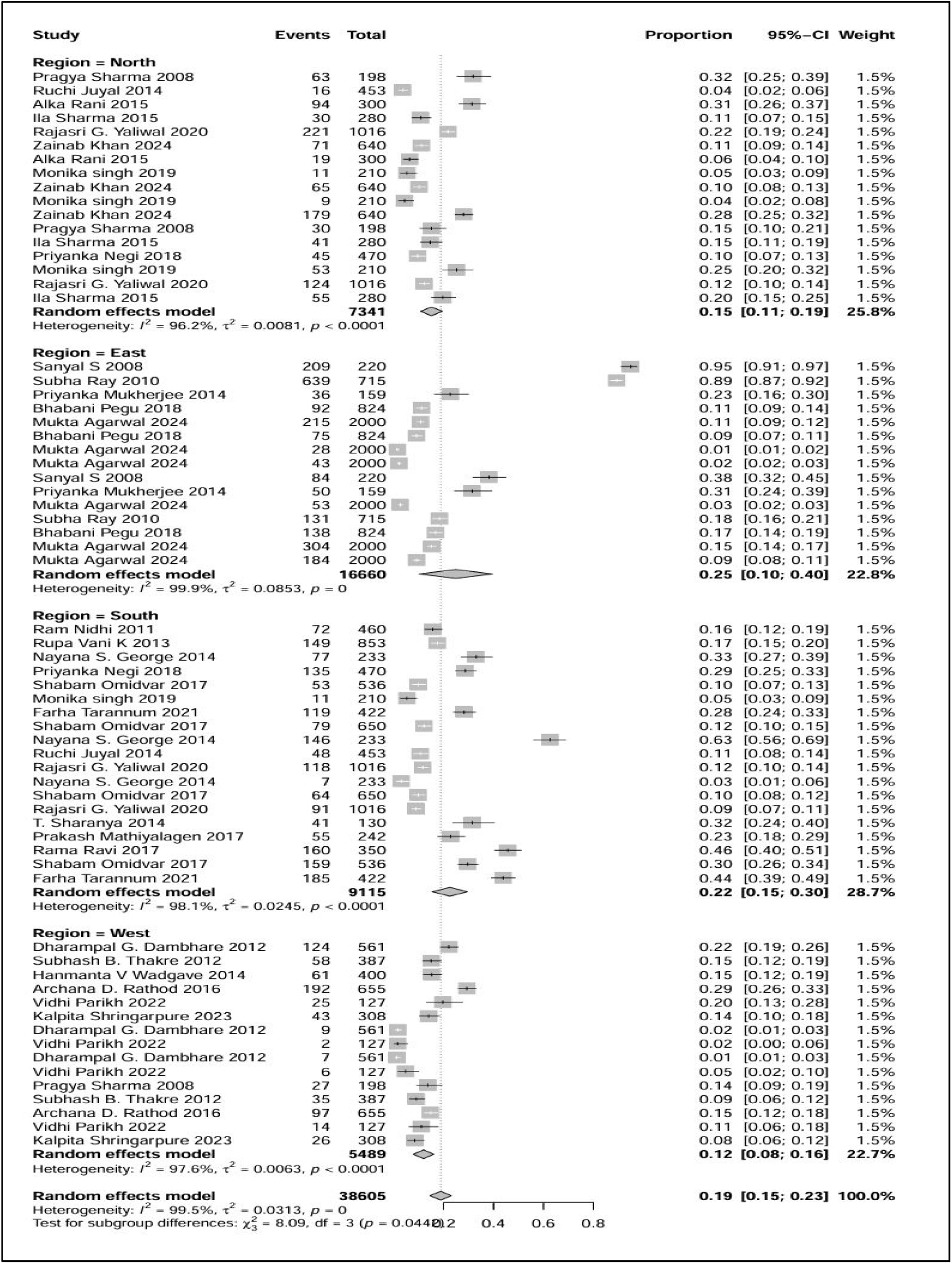
Sub-group –analysis by geographical region.

**Figure 7:**
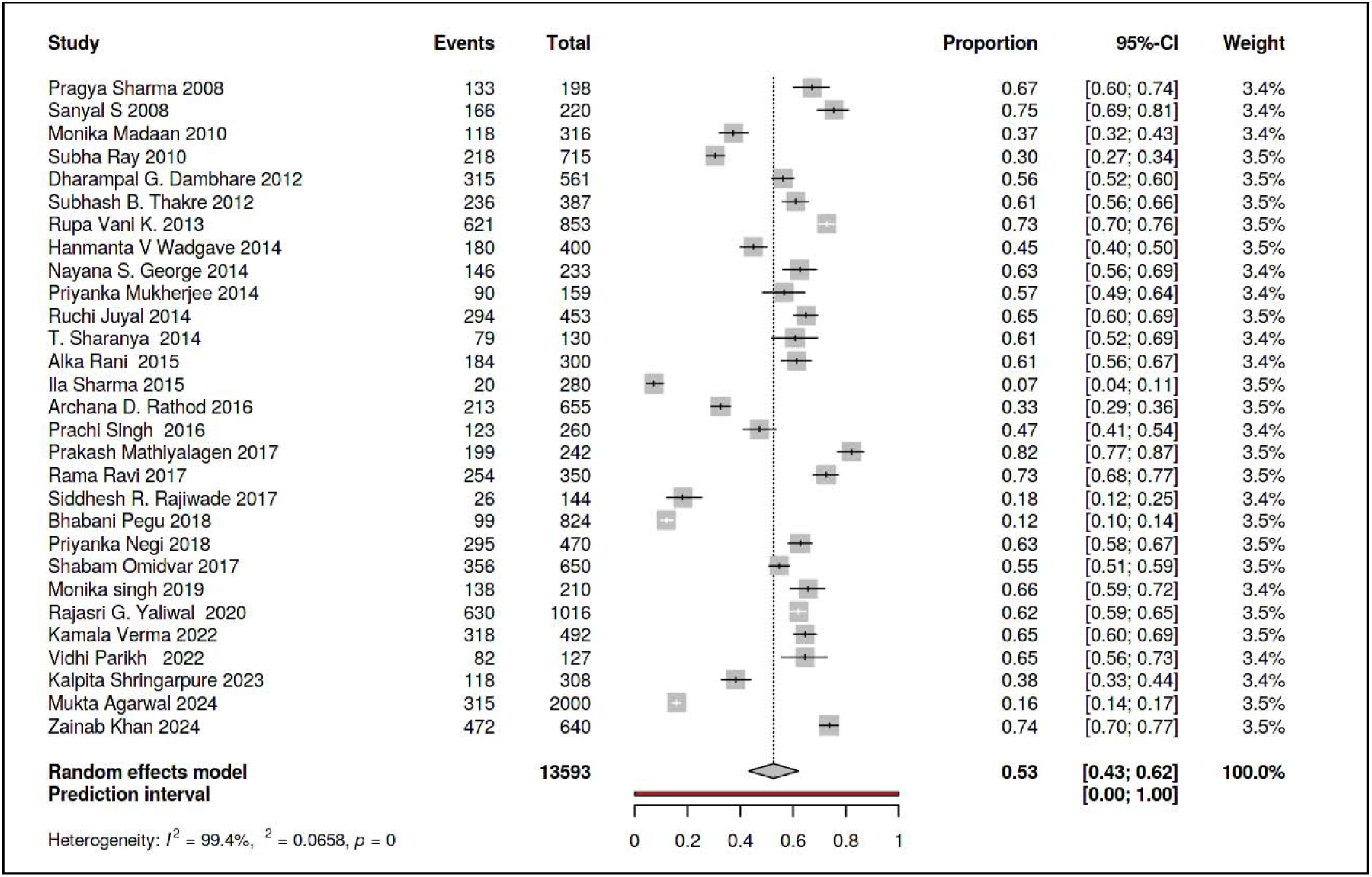
Forest plot showing pooled prevalence of Dysmenorrhoea among Adolescent girls in India.

### Burden due to commonest menstrual morbidity in adolescent girls of India: Dysmenorrhoea

The pooled prevalence of dysmenorrhoea was 53% (95% CI: 43-62%, I² = 93%), with individual study estimates ranging from 14.3% to 73.6%.

## Discussion

This systematic review and meta-analysis provide a comprehensive assessment of abnormal uterine bleeding (AUB) and its sub-types among Indian adolescent girls. The pooled prevalence of AUB in this study was found to be 19%, underscoring this condition as a significant public health issue in India. According to global studies, the prevalence of AUB ranges between 10% and 30%.(Liu et al., 2007) Notably, a systematic review conducted by Harlow indicated that the prevalence of AUB in developing countries falls between 10% and 15%, which aligns with our findings.(Harlow et al., 2025) Evaluating AUB has always posed challenges, primarily due to its complex nature, which encompasses a variety of symptoms and sub-types. The lack of standardized tools for assessment and the reliance on self-reported data from participants further complicate the evaluation process.

Our subgroup analysis of AUB sub-types sheds light on the epidemiological patterns within India, providing valuable insights for targeted screening and effective referral management. The three most prevalent AUB sub-types identified in this systematic review are oligomenorrhea (25%), metrorrhagia (24%), and menorrhagia (18%). The cultural silence and stigma surrounding menstrual health in India and other parts of the world contribute to the underreporting of these conditions, despite their association with various underlying health issues.(Castro & Czura, 2025) Addressing these cultural barriers is essential for improving awareness, encouraging open discussions, and facilitating better management of AUB among adolescents. By prioritizing education and support, we can enhance the detection and treatment of AUB, ultimately promoting better reproductive health outcomes for young women

In our study, we found the pooled estimate of oligomenorrhea to be 25%. In contrast, a systematic review conducted in Iran reported a pooled prevalence of only 13.1%, which is significantly lower than our findings. Additionally, a large-scale study focusing on an Italian adolescent population found the prevalence of oligomenorrhea to be just 3.4%. Interestingly, another study examining young athletes revealed a high prevalence of 24%, which closely aligns with our results. (Verrilli et al., 2018) Oligomenorrhea, characterized by infrequent menstrual periods, is a significant gynecological issue that can lead to considerable anxiety among students and those around them. Beyond the emotional impact, oligomenorrhea is associated with various health risks, including infertility, cardiovascular complications, and metabolic disturbances. Conditions such as obesity, hyperandrogenism, and metabolic syndrome are linked to this menstrual irregularity, highlighting the importance of addressing oligomenorrhea not only as a reproductive health concern but also as a broader public health issue.(Glueck et al., 2015; He et al., 2020)

Our study reported a pooled prevalence estimate of 25% for menorrhagia and 15% for hypermenorrhea. In contrast, a systematic review and meta-analysis conducted in Iran found the pooled prevalence of menorrhagia to be 13.1% and hypermenorrhea to be 12.4%.(Omani Samani et al., 2018) Additionally, a study conducted in Canada among adult and adolescent women revealed a pooled estimate of heavy menstrual bleeding (HMB) at 30%.(Liu et al., 2007) It is important to note that HMB is defined as heavy bleeding exceeding 80 ml and encompasses both menorrhagia and hypermenorrhea. Interestingly, a cohort study from the Netherlands reported a much lower incidence rate of HMB, highlighting regional variations in prevalence.(van Hooff et al., 2004) Furthermore, another systematic review conducted across ten Asian countries indicated a significantly higher pooled estimate of 48.6% for HMB. (Sinharoy et al., 2023)

These findings collectively suggest a notable discrepancy in the prevalence of heavy menstrual bleeding between developing and developed nations. The higher prevalence rates observed in developing countries may reflect a lack of health-seeking behavior, limited access to healthcare resources, and the absence of standardized screening tools for menstrual disorders. Cultural stigma and insufficient education surrounding menstrual health further hinder the proper management and referral of these conditions. By addressing these barriers, we can improve the identification and treatment of menstrual disorders, ultimately enhancing the quality of life for affected individuals. Overall, this results comparison across many studies highlights the need for increased awareness, education, and the implementation of standardized screening practices in developing nations like India. Other systematic review and meta-analysis revealed the three most common etiologies for HMB are von willebrand disease, platelet function defect and thyroid disease further highlighting the severity of this problem. (Comishen et al., 2025)

Menstrual problems include dysmenorrhea, premenstrual symptoms, menorrhagia, polymenorrhea, abnormal vaginal bleeding, amenorrhoea, oligomenorrhea, and irregular menstruation and they are the major source of absenteeism and poor academic performance in young women. Rafique et al. observed that menstrual problems such menorrhagia, irregular uterine bleeding, and polymenorrhea account for over 12% of gynecology referrals and have a high surgical intervention rate among young females.(Rafique & Al-Sheikh, 2018) Among the menstrual problems, dysmenorrhea is a major health issue for women worldwide. Dysmenorrhea is a prevalent ailment among adolescent girls, impacting quality of life, work productivity, and healthcare utilization across all reproductive years.(Molla et al., 2022) A recent systematic review done in Indian women reported the prevalence of dysmenorrhea ranging from 46% to 76% which was coinciding with our study findings, pooled prevalence of dysmenorrhoea was 53% (95% CI: 43-62%, I² = 93%).(Das & Jungari, 2024) In contrast a higher pooled prevalence estimate of dysmenorrhea was observed among the female students in Ethiopia; 71.69% (95% confidence interval (CI) = 66.82–76.56%) with a significant heterogeneity showing I^2^ = 94.2% and p < 0.00.(Molla et al., 2022) It can be attributed to several sociodemographic, cultural, and methodological factors. One of the primary reasons could be the difference in the study populations. Differences in awareness levels and health-seeking behavior, including access to menstrual health education, might have also influenced these findings. Another systematic review and meta-analysis done in Iran estimated a combined prevalence of primary dysmenorrhea as 73.27% (95% CI=65,12%-81.42%) which was also above than our findings. Their analysis showed considerable heterogeneity among studies (Q=4097.93, d.f.=25, p<0.001 and I^2^=99.4%). An individual study conducted in Ghaziabad district among women of 15-45 years of age residing in urban area found the prevalence of primary dysmenorrhea as 67.8% which was aligning with our study results.(Verma et al., 2024)

Polymenorrhea is a menstrual irregularity in which the menstrual cycles occurs at a higher frequency than normal i.e. less than 21 days.(Omani Samani et al., 2018) A recent systematic review done among the Indian women estimated the prevalence of polymenorrhea as 3–22.2% which was aligning with our findings which estimated a pooled prevalence as 7% (95% CI: 4-11%, range: 1-12%).(Das & Jungari, 2024) A study conducted in Kerala, estimated the prevalence as 3.1% and a systematic review done in developing countries among women in India self-reported a prevalence of polymenorrhea as 1% while 6% of women in Turkey complained of having frequent periods similarly, a lower prevalence of polymenorrhea was observed in most of the previous literature.(Harlow & Campbell, 2004)

Hypomenorrhea is a disorder characterized by light uterine bleeding i.e. in terms of either duration or days(<2 days) or both.(Kulshrestha & Durrani, 2019) Our study estimated a pooled prevalence of 12% (95% CI: 4-21%), with high heterogeneity (I²=97.2%, τ²=0.0164, p<0.0001)among Indian adolescent girls. The study findings of Das et al. with the prevalence observed as 5.1% in an individual study was lying within our range.(Das & Jungari, 2024) The prevalence of hypomenorrhea in Iran, as reported in the systematic review, was 5.25% (95% CI: 3.20%-7.30%), with estimates ranging from 0.9% to 12.90% across different studies included in the analysis.(Omani Samani et al., 2018) The literature indicates that hypomenorrhea is often under-recognized, particularly in adolescents, where its identification is not routinely practiced despite its potential implications for menstrual health.(de Sanctis et al., 2022)

A systematic review and meta-analysis conducted in Iran reported a pooled prevalence of metrorrhagia at 6.04% (95% CI: 1.99–10.08%), which is considerably lower than the 24% estimated in our study.(Omani Samani et al., 2018) In contrast, a cross-sectional study conducted among secondary school girls in Nigeria found a higher prevalence of 18.6% which is closer to our findings and may reflect similar adolescent-focused populations and settings.(U C Igbokwe And et al., 2021). Differences in the study population could be the cause for variation in study findings as in Iranian meta-analysis included a wider age range than our study, which focused primarily on adolescents, who are more prone to anovulatory cycles. There are only few Indian meta-analyses which offers pooled prevalence for metrorrhagia among adolescents while larger reviews and individual studies show significant heterogeneity. In a two-decade assessment of menstruation diseases in India, prevalence ranged from 3% to 87%, indicating variation in classifications, settings, and demographics.(Das & Jungari, 2024) A study in Puducherry found metrorrhagia in 1.1% of adolescents with menstrual disorders.(Rajiwade et al., 2018) These results show that our study’s pooled estimate of 24% (95% CI: 4–44%), though higher than clinical estimates, is compatible with community-level data, especially when broader menstrual abnormalities, self-reports, and adolescent-specific variables are included.

## Limitations

There are certain limitations observed with this review. Firstly,high statistical heterogeneity (I² > 90%) observed across all analysis limits the reliability and generalization of pooled estimates of AUB and its sub-types on Indian adolescents girls. Second limitation is lack of use of standardized method or tool or diagnostic criteria for data collection, which can directly associated with pooled estimates. Additionally, biases attributed due to language restriction , searches limited to peer-reviewed journal, variation in study settings and method of data collection has been not explored in this study, which may cover up real burden due to AUB among adolescents girls.

## Conclusion

This systematic review and meta-analysis provided strong national-level pooled estimates of abnormal uterine bleeding and its sub-types in India. This study found that oligomenorrhea, metrorrhagia, and menorrhagia are the most common AUB sub-types among adolescent girls in India. Dysmenorrhea affects more than half of teenage girls. The results necessitate enhanced menstrual health systems, encompassing regular screening with standard tool, prompt diagnosis, focused therapies, and proactive care to avert enduring problems. Significant variability among studies, influenced by regional and methodological disparities, highlighted the critical necessity for consistent definitions and diagnostic criteria in forthcoming research to enhance the alignment of prevalence estimates, clinical care, and policy.

## Data Availability Statement

All data analyzed in this study are available from publicly accessible sources. Additional data supporting the findings of this study are available from the corresponding author upon reasonable request.

## Ethics Approval Statement

As this study is a systematic review and does not involve human or animal subjects, ethics approval was not required.

